# Sharp rises in alcohol-induced deaths in the United States (1999-2022) across genders, ages, races

**DOI:** 10.1101/2024.12.10.24318758

**Authors:** Tony Wong, Lucas Böttcher, Tom Chou, Maria R. D’Orsogna

## Abstract

**Importance:** Alcohol-induced deaths rose by 163% in the United States between 1999 and 2022, with sharp increases across most gender, age, and race groups in Spring 2020, concurrent with the onset of COVID-19. This study analyzes monthly mortality rates by demographics from 2018 to 2022.

**Objective:** To determine which demographic groups in the United States experienced the largest variations in alcohol-induced crude rates, yearly between 1999 and 2022, and monthly between 2018 and 2022.

**Design, setting, and participants:** This cross-sectional study used yearly (1999 to 2022) and monthly (January 2018 to December 2022) mortality data from the Centers for Disease Control and Prevention’s Wide-Ranging Online Data for Epidemiologic Research. Fourteen alcohol-related causes of death were identified according to the International Statistical Classification of Diseases and Related Health Problems, Tenth Revision. Population data was used to determine crude rates for given demographic groups.

**Main outcomes and measures:** The month and magnitude of sharp rises in crude rates were determined using Bayesian regression implemented through Rbeast. Data were stratified by gender, age, race and/or three classes of cause-of-death (alcohol liver disease; mental and behavioral disorders due to alcohol use; alcohol poisoning).

**Results:** Alcohol-induced deaths for individuals aged 15 and older increased 163% from 19,458 in 1999 to 51,184 in 2022, with a peak in 2021. Most affected were those aged 25-34, with a +345% (males) and a +441% (females) increase from 1999 to 2022. Rbeast applied to monthly data shows that for most gender, age, race groups, mortality rose sharply in Spring 2020 due to alcohol-related liver disease and mental and behavioral disorders. The largest relative increases by race were among American Indian and Alaska Native males with a 42% jump (May-Jun ‘20) and Black females with a 33% jump (Apr-May ‘20). By age, those between 35–44 (both genders) experienced the largest increase with a 29% jump (Apr-May ‘20).

**Conclusions and relevance:** Alcohol-induced deaths have been steadily increasing since 1999, with major increases emerging in Spring 2020 for most gender, age, race groups. Policies to reduce excessive alcohol consumption, address the social determinants of addiction, and improve access to treatment can be most effective when targeted to specific groups.

**Key Points:** *Question:* How do trends in alcohol-induced deaths vary by gender, age, race, and specific causes of death between 1999 and 2022 in the United States?

*Findings:* From 1999 to 2022, alcohol-induced deaths surged across most demographics, with female crude rates increasing faster than males. Mortality spiked in Spring 2020, concurrent with the start of the COVID-19 pandemic, with the largest relative increases observed among American Indian and Alaska Native males (+42%) and Black females (+33%).

*Meaning:* Targeted prevention and intervention efforts are crucial for addressing differential trends in alcohol-induced deaths, which were likely worsened by social isolation and treatment disruptions during the pandemic.

## Introduction

Alcohol use disorder (AUD) continues to pose a significant public health challenge in the United States. Excessive alcohol consumption causes severe complications, such as alcoholic liver disease [1, 2, 3, 4], and contributes to metabolic syndrome, an ensemble of co-occurring conditions such as arterial hypertension, diabetes or obesity or other chronic diseases [5, 6, 7, 8]. The problem affects both genders [7, 8, 9], and has been exacerbated by the increasing popularity of binge drinking [10, 11, 12] and the practice of mixing alcohol with medications or illicit substances [13]. Even moderate consumption can be a risk factor for older adults with health issues [14].

To track the long-term dynamics of alcohol-induced deaths in the United States, we normalized age- and race-stratified yearly crude rates between 1999 and 2022, relative to their 1999 values. Although the overall number of female deaths is lower than that of males, crude rates among females are increasing at a much higher rate than among males, for all ages and races, as shown in Figure 1. Age stratification reveals that the largest increase in mortality for both genders arise in the 25-34 age group, and that crude rates peaked in 2021 for all ages. Race stratification was possible only until 2020 when Centers for Disease Control and Prevention’s Wide-Ranging Online Data for Epidemiologic Research (CDC WONDER) changed classification criteria.

**Figure 1.**
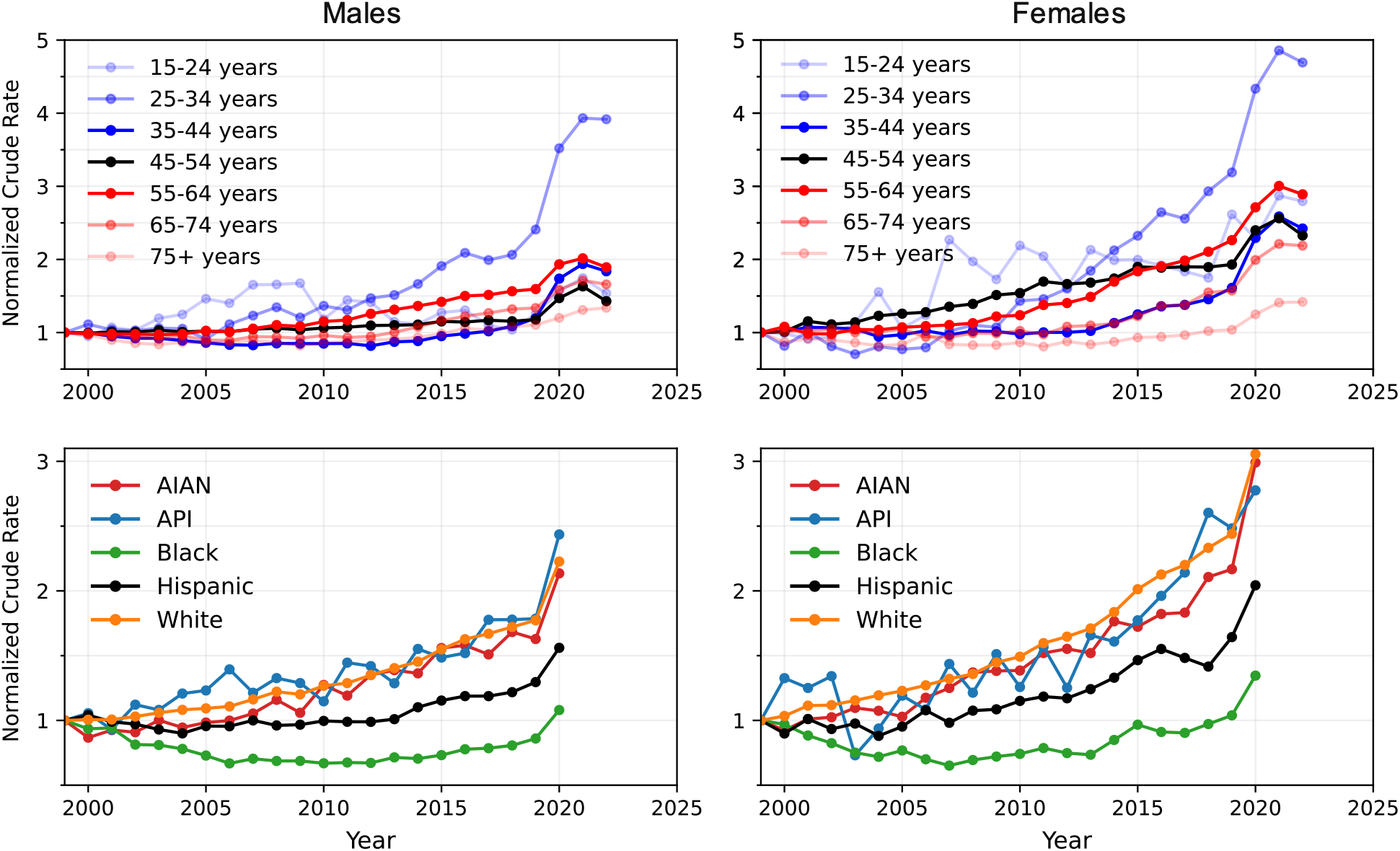
Yearly crude rates normalized by their 1999 values. Normalized crude mortality rates for males (left) and females (right), stratified by age (top) from 1999 to 2022, and race (bottom) from 1999 to 2020. “Bridged race” categories are used. Since racial categorizations in the CDC WONDER databases were changed in 2020 to “single race 6” categories, race analyses that started in 1999 cannot extend beyond 2020. Values below 1 indicate crude rate decreases relative to their 1999 value, those above 1 indicate increases. Non-normalized crude rates are plotted in eFigure 1 of the Supplement.

Between 1999 and 2020, the largest increase in crude rates is observed for the American Indian and Alaska Native (AIAN), Asian and Pacific Islander (API), and White populations. Crude rates increased at a much slower rate among Hispanics. Remarkably, alcohol-induced mortality in the Black population decreased until the mid-2010s and crude rates remained below their 1999 values until 2019, rising again in 2020. Overall, steep upward trends are observed in recent years for most demographic groups. Building on reports of accelerated alcohol-related deaths during the COVID-19 pandemic [15, 16, 17], we conduct a detailed monthly trend analysis to examine how increases in alcohol-induced mortality are distributed across demographic groups between 2018 and 2022.

## Methods

### Data source

We analyze mortality data for US individuals aged 15 and older from 1999 to 2022, as sourced from the CDC WONDER database [18]. Each recorded death has a single underlying cause, with up to twenty additional contributing causes. Alcohol-related deaths are identified using specific codes from the International Statistical Classification of Diseases and Related Health Problems, Tenth Revision (ICD-10). These include K70 (alcoholic liver disease), F10 (mental and behavioral disorders due to the use of alcohol), and four codes for alcohol poisoning, R78, X45, X65, Y15. Alcoholic liver disease (ALD), mental and behavioral disorders due to the use of alcohol, and alcohol poisoning are the three main categories of alcohol-induced deaths. Other alcohol-related causes of death are listed under E24.4, G31.2, G62.1, G72.1, I42.6, K29.2, K85.2, and K86.

We follow the racial categorization provided by the CDC WONDER database, which uses “bridged race” categories (White, Black, AIAN, API) for mortality data between 1999 and 2020, and “single race 6” categories (White, Black, AIAN, Asian, Native Hawaiian or other Pacific Islander, Mixed race) for mortality data from 2018 onwards [18]. While both classifications are available for 2018-2020, single race 6 is adopted exclusively from 2020 onwards. Thus, meaningful racial comparisons can extend only from 1999 to 2020 or from 2018 onwards. To derive fatalities for the Hispanic population we select “All races” and “Hispanic or Latino” origin; to derive fatalities for all other races we select the race of interest and “Not Hispanic or Latino” origin.

Yearly data (used in Figure 1) is based on bridged race categories; monthly data (used in Figure 2 and 3) is based on single race 6 categories. We utilize 10-year age groups, merging the 15-24 and 24-35 age populations for the monthly data due to small numbers of fatalities. For each demographic group, the yearly (monthly) crude rate is calculated as the number of fatalities in each year (month) divided by the associated total population in that given year (month), multiplied by 100,000. Yearly population estimates are made available by the US Census Bureau and included in the CDC WONDER database. Monthly population estimates are obtained from the online database on the US Census Bureau’s website [19, 20].

**Figure 2.**
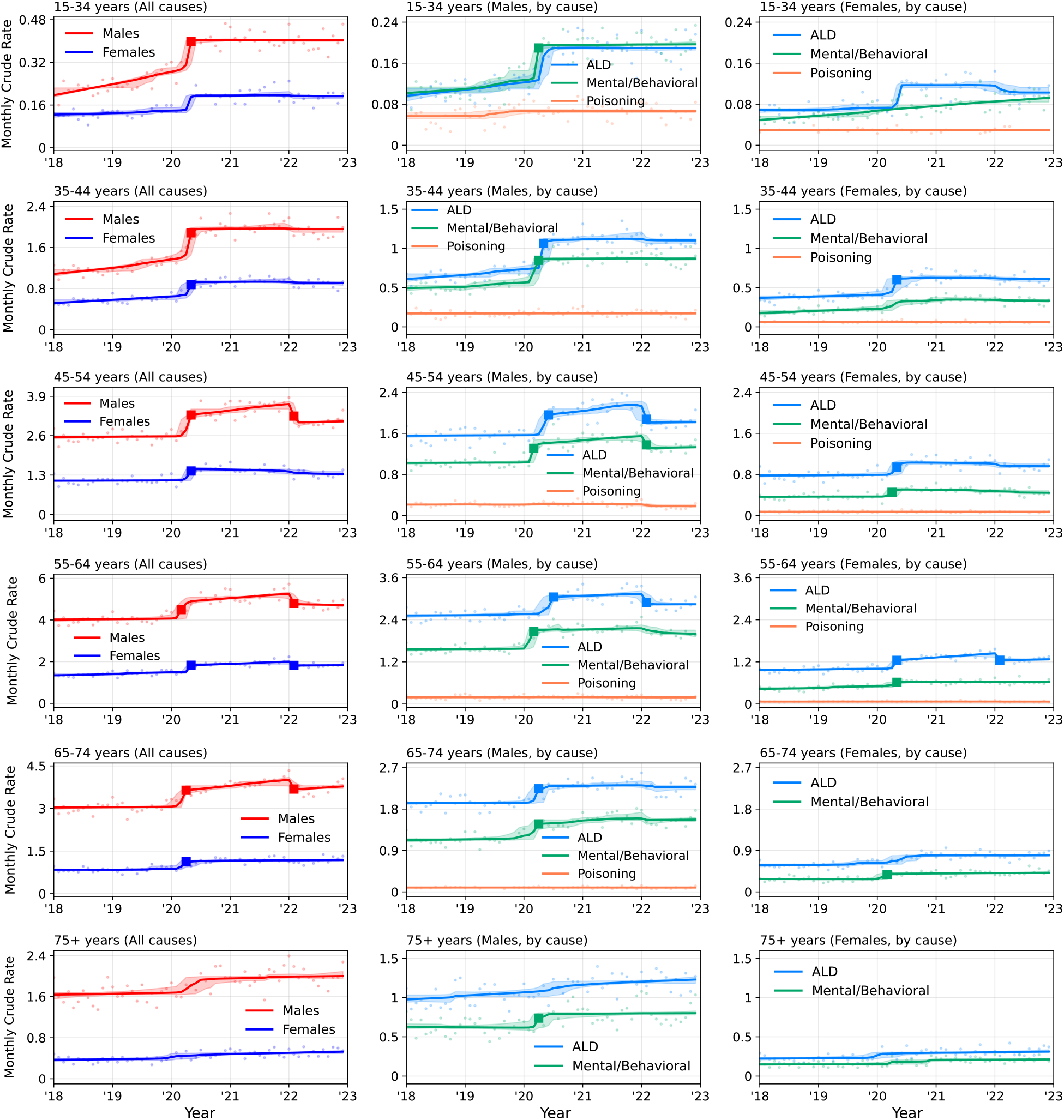
Alcohol-related monthly crude rates between 2018 and 2022, stratified by gender, age (first column) and by gender, age and cause of death (second, third columns). Data are indicated as dots, solid curves represent Rbeast-derived trends, shaded areas their CIs, and solid squares indicate statistically significant TCP jumps. Except for those aged 75 or over, all ages experienced a statistically significant TCP jump in the Spring of 2020. Males and females aged 55-64 display the largest mortality, but the largest relative increase was among males aged 15-34 (+28%, Apr-May ‘20) and 35-44 (+29%, Apr-May ‘20) and females aged 35-54 (+29%, Apr-May ‘20). TCP jump values are listed in eTable 1 and eTable 2 of the Supplement.

**Figure 3.**
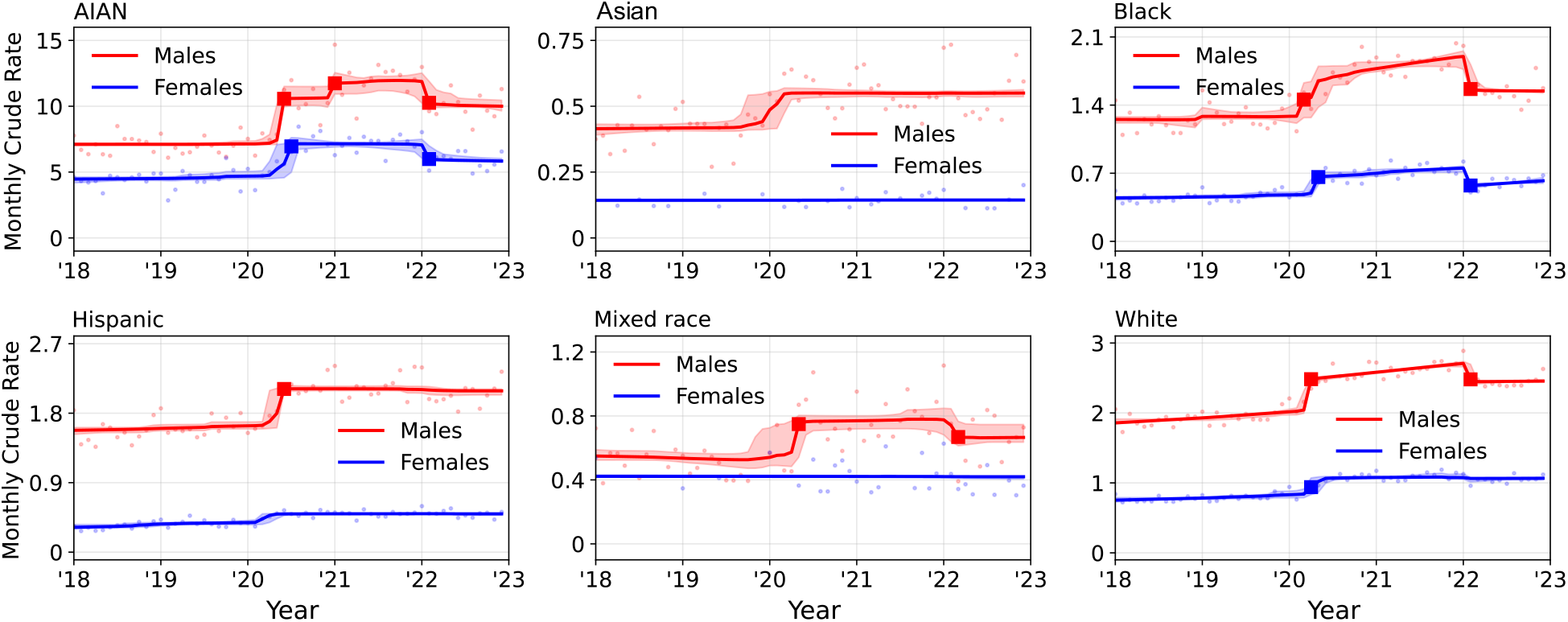
Alcohol-related monthly crude rates for ages 15 and older between 2018 and 2022, stratified by gender and race (using the single race 6 classification). Data are indicated as dots, solid curves represent Rbeast-derived trends, shaded areas represent their CIs, and solid squares mark statistically significant TCP jumps. Except for Asians, males of all races experience a TCP jump in the Spring of 2020 as follows: AIANs: +42% males (May-Jun ‘20) and +24% females (Jun-Jul ‘20); Blacks: +13% males (Feb-Mar ‘20) and +33% females (Apr-May ‘20); Hispanics: +18% males (May-Jun ‘20); Mixed race +31% males (Apr-May ‘20); Whites: +22% males (Mar-Apr ‘20) and +12% females (Mar-Apr ‘20). Male trends are much higher than female trends, and never return to pre-pandemic levels.

### Statistical analysis

We apply the Bayesian regression software Rbeast [21] to monthly crude rates between January 2018 and December 2022 for trend analysis. Rbeast extracts the trend and seasonal components of a (possibly noisy) time series and detects trend change points (TCPs) characterized by abrupt jumps between two consecutive trend curves across a single data point. Rbeast also provides 95% credible intervals for the trend segments and TCP locations. We utilize Rbeast to capture the trend in monthly crude rates, and to exclude seasonal effects and statistical fluctuations.

Mathematically, Rbeast outputs the trend function *T*(*t*_*i*_) which describes the non-seasonal, deterministic component of the appropriate mortality at time point *t*_*i*_. The trend function is comprised of a probability-weighted average of component trend functions, each describing a different statistical model with different sets of piecewise linear segments. The TCP is defined as TCP(*t*_*i*_)≡*T*(*t*_*i*_)−*T*(*t*_*i*−1_), the change in trend between its values at *t*_*i*_ and *t*_*i*−1_, and allows one to identify jumps or significant discontinuities in the trend. For a TCP to be classified as an abrupt jump within month *t*_*i*_, we require that the relative TCP, TCP(*t*_*i*_) ⁄*T*(*t*_*i*−1_), exceeds 0.05. Trend values before and after a TCP, *T*(*t*_*i*−1_) and *T*(*t*_*i*_), for all demographic groups, are shown in Figures 2 and 3 and listed in eTables 1, 2, and 3 of the Supplement.

## Results

We focus on alcohol-induced deaths between 2018 and 2022 for individuals aged 15 and older, using single race 6 categories. Rbeast applied to monthly crude rates reveals statistically significant, abrupt upward TCP jumps in mortality trends in Spring 2020, concurrent with the onset of COVID-19, for both genders and all age groups below 75 years. As shown in Figure 2, the largest TCP jumps occurred in younger cohorts, with male mortality trends increasing by 28% for ages 15-34 between April and May 2020, and by 29% for ages 35-44 in the same period. Among females, the largest TCP jump was for those aged 35-44 whose mortality trend rose by 29% between April and May 2020. For both genders and all age groups less than 75 years, the trend never returns to pre-pandemic levels, despite some groups experiencing downward TCP jumps, suggesting long-lasting effects.

Further stratifications of monthly crude rates by cause of alcohol-induced death are also shown in Figure 2 for both genders. Alcoholic liver disease (ALD) emerges as the main cause of death, followed by mental and behavioral disorders due to alcohol use, especially for younger cohorts. Alcohol poisoning remains uniformly low and is not associated to any TCP jump. Among females, the upward TCP jumps for ALD are of the same magnitude as for all alcohol-induced deaths, implying that the mortality surge observed in Spring 2020 is mostly attributable to ALD, rather than to mental or behavioral disorders. Conversely, males aged 35-44 have a TCP jump in May ‘20 (change: 0.42) that aligns with TCP jumps in ALD (May ‘20, change: 0.32) and mental and behavioral disorders (Apr ‘20, change: 0.15). Stratifications by cause of death reveal that mortality due to alcohol-induced mental and behavioral disorders increased by 45% among males aged 15-34 in one month (Mar-Apr ‘20). ALD deaths rose by 42% for males aged 35-44, and by 34% for females aged 35-44 in the same month (Apr-May ‘20). Figure 2 also shows that deaths due to alcohol-induced mental and behavioral disorders among females aged 15-34 have been steadily increasing since 2018.

Results stratified by race are shown in Figure 3. Except for Asians, males of all races experienced significant upward TCP jumps in Spring 2020, concurrent with the onset of the COVID-19 pandemic. AIAN males suffer from disproportionately high alcohol-induced mortality compared to other races even pre-pandemic since their pre-TCP monthly crude rate trend value (7.4, May ‘20) is four times higher than the next largest one (1.8, May ‘20) for Hispanics. AIAN males also experience the highest TCP jump in the Spring of 2020, with a post-TCP trend value of 10.6 (Jun ‘20), corresponding to a 42% rise in one month, much higher than that of other races. The further upward (Jan ‘21) and downward (Feb ‘22) TCP jumps for AIAN males keep the mortality trend higher than during pre-pandemic times. AIAN females also have the highest pre-TCP monthly crude rate trend value (5.6, Jun ‘20), but, surprisingly, the highest relative TCP jump is for Black females, whose mortality trend increased by 33% between April and May 2020. For comparison, the TCP jump among Black males is 13% between February and March 2020.

## Discussion

### Limitations

Underlying cause of deaths are selected from a list of alcohol-induced causes, as itemized by the CDC WONDER database. Deaths due to chronic diseases related to alcohol use were not included, potentially underestimating the overall death burden. Since the database suppresses entries with fewer than 10 deaths, data is not available for combinations of gender, age or races with extremely low fatalities. This is the case for monthly mortality data for those aged 15-24 between 2018 to 2022. We thus combined the 15-24 and 25-34 age groups into a single 15-34 category to ensure sufficient data for the Rbeast trend analysis. We did not analyze the data geographically although quantifying state or county variations in alcohol-induced deaths could be useful for a more detailed view and for forecasting regional trends [22].

## Conclusions

Alcohol-induced mortality is one of the major contributors to the “deaths of despair” crisis in the United States, in addition to drug-induced mortality and suicide, affecting many socioeconomic and demographic groups [23, 24, 25]. We analyzed the steady rise of yearly crude rates for alcohol-related fatalities between 1999 and 2022 and the abrupt rise that occurred in 2020 for all demographics. Yearly trends between 1999 and 2022 reveal that although mortality is highest among men, crude rates are increasing faster among females, across all ages and races. Similarly, although mortality is highest among those aged 55-64 for both genders, the largest increases were for those aged 25-34. For females in this age group, crude rates increased almost five-fold between 1999 and 2022; for males they increased almost four-fold.

The use of Rbeast allows us to identify changes in trends in the monthly crude mortality rates within age, race, gender groups, and causes of death. For all demographic groups, the month over which crude mortality rates jumped were between March and June 2020, near the start of COVID-19 pandemic. The most affected were AIAN males and Black females, whose monthly crude rates rose by 42% and 33% in one month, respectively, and males and females aged 35-44, whose mortality both rose by 29% in one month. These abrupt rises are primarily linked to ALD (especially for women) and mental and behavioral disorders, with no notable increase in alcohol poisoning for either gender.

A worthwhile direction for future work would be to examine whether, and to what degree, social isolation and disruptions in AUD treatment imparted by COVID-19 may have contributed to the large spikes in monthly mortality crude rates observed in the Spring of 2020. While most mortality trends began decreasing or stabilizing in early 2022, they remain elevated compared to pre-pandemic levels, suggesting long-lasting impacts.

The rise in alcohol-induced mortality, and the heterogeneous trends across demographics, highlight the need for targeted prevention and treatment efforts, particularly for males, youth, and the AIAN population. Although mortality is lower, the accelerated rise in crude rates among females relative to males is a major cause for concern. Intervention is particularly urgent given that elevated mortality is only one of many negative outcomes associated with excessive alcohol consumption, a crisis which impacts public health, safety, healthcare expenditures, and family stability.

## Data Availability

All data used in this study are publicly available at https://wonder.cdc.gov/.

