## Supplemental Material for "Sharp rises in alcohol-induced deaths in the United States (1999-2022) across genders, ages, races"

**eMethods.**

**eFigure 1.** Yearly crude rates stratified by age from 1999 to 2022 and by race from 1999 to 2020

**eTable 1.** TCP jumps for male monthly crude rates stratified by age group and cause of death from 2018 to 2022

**eTable 2.** TCP jumps for female monthly crude rates stratified by age group and cause of death from 2018 to 2022

**eTable 3.** TCP jumps for monthly crude rates stratified by races for both genders from 2018 to 2022

This supplemental material has been provided by the authors to give readers additional information about their work.

### eMethods.

#### Yearly crude rates

In eFigure 1, we plot the non-normalized yearly crude rates stratified by age (between 1999 and 2022) and by race (between 1999 and 2020). The normalized crude rates plotted in Figure 1 of the main paper are given by the yearly values in eFigure 1 divided by the corresponding 1999 entry. Males are disproportionally affected in all groups. Age-wise, the most affected are those between 55-64 for both genders; for both males and females, mortality roughly doubled in this age group between 1999 and 2022. Race-wise, the AIAN population displays the highest crude rates for both genders; for both males and females, the mortality more than doubled between 1999 and 2020.

#### Statistically significant TCP jumps, 2018-2022

In eTables 2 and 3, we show the values of the statistically significant trend change point (TCP) jumps obtained from Rbeast applied to monthly crude rates. The quantity $T(t_i)$ represents the Rbeast-derived trend at month $t_i$. The TCP is defined as TCP$(t_i) \equiv T(t_i) - T(t_{i-1})$. For a TCP to be classified as an abrupt jump at month $t_i$, we require that the relative TCP, TCP$(t_i)\ /T(t_{i-1})$, exceeds 0.05. Trend values before and after a TCP, $T(t_{i-1})$ and $T(t_i)$, for all demographic groups are listed in eTables 1, 2, and 3 and are shown in Figures 2 and 3 of the main text.

**eFigure1. Yearly crude rates stratified by age from 1999 to 2022 and by race from 1999 to 2020**

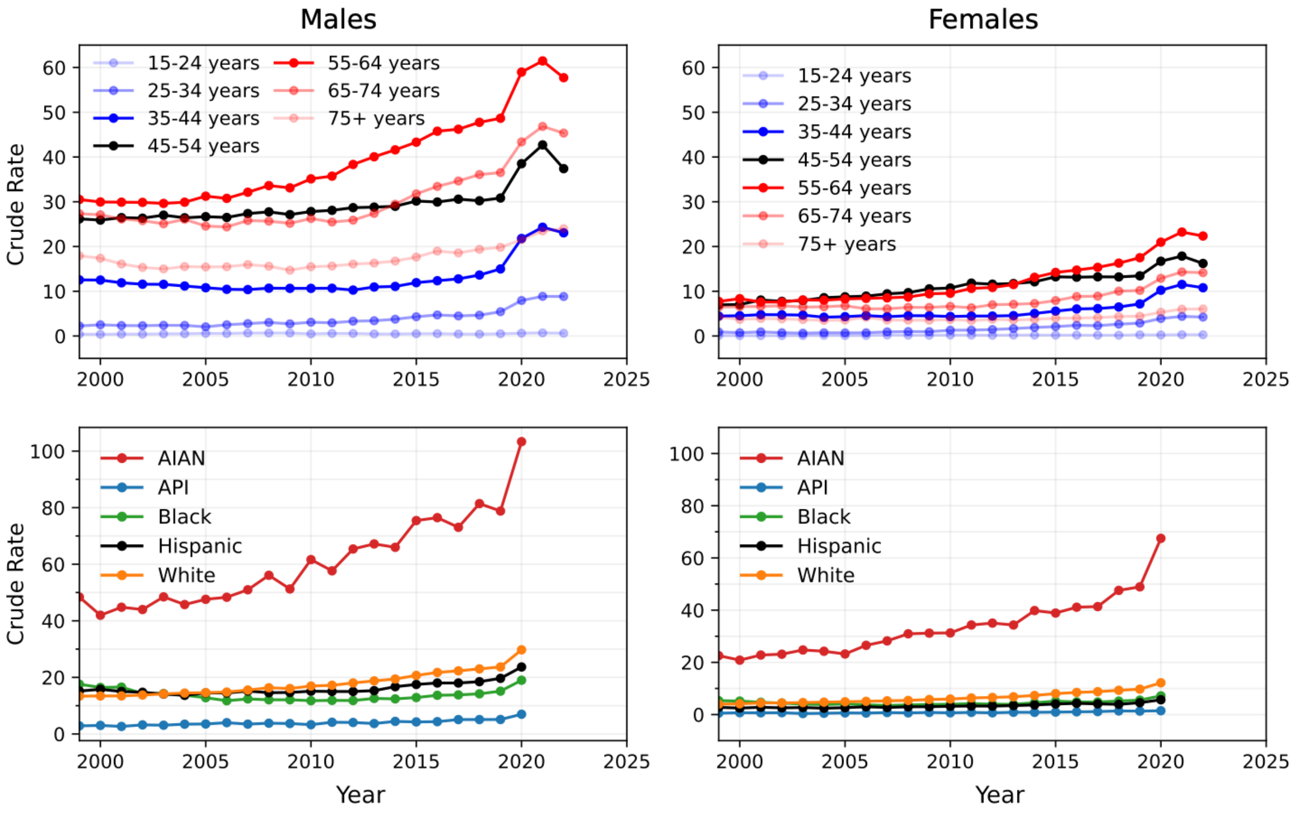


The yearly crude rates for males (left) and females (right), stratified by age (top) from 1999 to 2022 and by race (bottom) from 1999 to 2020. Yearly crude rates for a given demographic are defined as the number of fatalities of that demographic in a given year, divided by the entire demographic population, multiplied by 100,000. These crude rates were used to create Figure 1 in the main text.

**eTable 1: TCP jumps for male monthly crude rates stratified by age group and cause of death from 2018 to 2022**

| Age Group | Cause | Trend $T(t_{i-1})$ | Trend $T(t_i)$ | TCP Jump (pct chg) | Credible Interval |
|---|---|---|---|---|---|
| 15-34 | All causes | $T$(Apr '20) = 0.31 | $T$(May '20) = 0.40 | May '20 (+28%) | Mar-Jun '20 |
| 15-34 | Mental/ Behavioral | $T$(Mar '20) = 0.13 | $T$(Apr '20) = 0.19 | Apr '20 (+45%) | Mar-Jun '20 |
| 35-44 | All causes | $T$(Apr '20) = 1.47 | $T$(May '20) = 1.89 | May '20 (+29%) | Mar-Jul '20 |
| 35-44 | ALD | $T$(Apr '20) = 0.75 | $T$(May '20) = 1.07 | May '20 (+42%) | Apr-Jul '20 |
| 35-44 | Mental/ Behavioral | $T$(Mar '20) = 0.70 | $T$(Apr '20) = 0.85 | Apr '20 (+21%) | Feb-Jun '20 |
| 45-54 | All causes | $T$(Apr '20) = 2.79 | $T$(May '20) = 3.29 | May '20 (+18%) | Mar-Jun '20 |
| 45-54 | All causes | $T$(Jan '22) = 3.65 | $T$(Feb '22) = 3.24 | Feb '22 (-11%) | Jan-Apr '22 |
| 45-54 | ALD | $T$(May '20) = 1.75 | $T$(Jun '20) = 1.96 | Jun '20 (+12%) | Apr-Jul '20 |
| 45-54 | ALD | $T$(Jan '22) = 2.13 | $T$(Feb '22) = 1.88 | Feb '22 (-12%) | Dec '21 - Apr '22 |
| 45-54 | Mental/ Behavioral | $T$(Feb '20) = 1.04 | $T$(Mar '20) = 1.31 | Mar '20 (+26%) | Feb-May '20 |
| 45-54 | Mental/ Behavioral | $T$(Jan '22) = 1.55 | $T$(Feb '22) = 1.38 | Feb '22 (-11%) | Jan-Apr '22 |
| 55-64 | All causes | $T$(Feb '20) = 4.09 | $T$(Mar '20) = 4.50 | Mar '20 (+10%) | Feb-Jun '20 |
| 55-64 | All causes | $T$(Jan '22) = 5.26 | $T$(Feb '22) = 4.80 | Feb '22 (-9%) | Jan-Apr '22 |
| 55-64 | ALD | $T$(Jun '20) = 2.76 | $T$(Jul '20) = 3.05 | Jul '20 (+10%) | Apr-Aug '20 |
| 55-64 | ALD | $T$(Jan '22) = 3.13 | $T$(Feb '22) = 2.90 | Feb '22 (-7%) | Jan-Apr '22 |
| 55-64 | Mental/ Behavioral | $T$(Feb '20) = 1.78 | $T$(Mar '20) = 2.07 | Mar '20 (+16%) | Jan-May '20 |
| 65-74 | All causes | $T$(Mar '20) = 3.26 | $T$(Apr '20) = 3.64 | Apr '20 (+12%) | Feb-May '20 |
| 65-74 | All causes | $T$(Jan '22) = 4.01 | $T$(Feb '22) = 3.68 | Feb '22 (-8%) | Jan-Mar '22 |

| **Age Group** | **Cause** | **Trend $T(t_{i-1})$** | **Trend $T(t_i)$** | **TCP Jump (pct chg)** | **Credible Interval** |
|---|---|---|---|---|---|
| 65-74 | ALD | $T$(Mar '20) = 1.99 | $T$(Apr '20) = 2.24 | Apr '20 (+13%) | Feb-Jun '20 |
| 65-74 | Mental/ Behavioral | $T$(Mar '20) = 1.32 | $T$(Apr '20) = 1.48 | Apr '20 (+12%) | Jun '19 - May '20 |
| 75+ | Mental/ Behavioral | $T$(Mar '20) = 0.63 | $T$(Apr '20) = 0.74 | Apr '20 (+17%) | Feb-Jul '20 |

Values are plotted in Figure 2 of the main text. The percent change (pct chg) at month $t_i$ is defined as 100 $\mathrm{TCP}(t_i)\ /T(t_{i-1})$, where $\mathrm{TCP}(t_i) = T(t_i) - T(t_{i-1})$ is the TCP jump. Mortality is highest among those between 55-64. The largest relative increase is for males aged 35-44 (TCP jump +29%, May '20) and 15-34 (TCP jump +28%, May '20). Deaths due to ALD rose the most for those aged 35-44 (+42%, May '20); deaths due to mental and behavioral issues rose the most for those aged 15-34 (+45%, Apr '20).

**eTable2. TCP jumps for female monthly crude rates stratified by age group and cause of death from 2018 to 2022**

| Age Group | Cause | Trend $T(t_{i-1})$ | Trend $T(t_i)$ | TCP Jump (pct chg) | Credible Interval |
|---|---|---|---|---|---|
| 35-44 | All causes | $T$(Apr '20) = 0.68 | $T$(May '20) = 0.88 | May '20 (+29%) | Mar-Jul '20 |
| 35-44 | ALD | $T$(Apr '20) = 0.45 | $T$(May '20) = 0.60 | May '20 (+34%) | Feb-Oct '20 |
| 45-54 | All causes | $T$(Apr '20) = 1.20 | $T$(May '20) = 1.44 | May '20 (+20%) | Mar-Jul '20 |
| 45-54 | ALD | $T$(Apr '20) = 0.83 | $T$(May '20) = 0.94 | May '20 (+14%) | Feb-Jul '20 |
| 45-54 | Mental/ Behavioral | $T$(Mar '20) = 0.38 | $T$(Apr '20) = 0.45 | Apr '20 (+19%) | Feb-Aug '20 |
| 55-64 | All causes | $T$(Apr '20) = 1.52 | $T$(May '20) = 1.83 | May '20 (+21%) | Apr-Jun '20 |
| 55-64 | All causes | $T$(Jan '22) = 2.00 | $T$(Feb '22) = 1.82 | Feb '22 (-9%) | Jan-Mar '22 |
| 55-64 | ALD | $T$(Apr '20) = 1.04 | $T$(May '20) = 1.25 | May '20 (+20%) | Mar-Jun '20 |
| 55-64 | ALD | $T$(Jan '22) = 1.44 | $T$(Feb '22) = 1.25 | Feb '22 (-14%) | Jan-Mar '22 |
| 55-64 | Mental/ Behavioral | $T$(Apr '20) = 0.55 | $T$(May '20) = 0.62 | May '20 (+13%) | Feb-Jul '20 |
| 65-74 | All causes | $T$(Mar '20) = 0.96 | $T$(Apr '20) = 1.13 | Apr '20 (+18%) | Feb-Jun '20 |
| 65-74 | Mental/ Behavioral | $T$(Feb '20) = 0.32 | $T$(Mar '20) = 0.38 | Mar '20 (+18%) | Dec '19 - May '20 |

Values are plotted in Figure 2 of the main text. The percent change (pct chg) at month $t_i$ is defined as 100 $\mathrm{TCP}(t_i)\ /T(t_{i-1})$, where $\mathrm{TCP}(t_i) = T(t_i) - T(t_{i-1})$ is the TCP jump. Mortality is highest among those between 55-64. The largest relative increase among females is for those aged 35-44 (TCP jump +29%, May '20). Deaths due to ALD rose the most for those aged 35-44 (+34%, May '20); deaths due to mental and behavioral issues rose the most for those aged 45-54 (+19%, Apr '20).

#### eTable3. TCP jumps for monthly crude rates stratified by races for both genders from 2018 to 2022

| Race | Gender | Trend $T(t_{i-1})$ | Trend $T(t_i)$ | TCP Jump (Pct chg) | Credible Interval |
|---|---|---|---|---|---|
| AIAN | Male | $T$(May '20) = 7.43 | $T$(Jun '20) = 10.57 | Jun '20 (+42%) | Apr-Jul '20 |
| AIAN | Male | $T$(Dec '20) =10.64 | $T$(Jan '21) = 11.75 | Jan '21 (+10%) | Dec '20 - Apr '21 |
| AIAN | Male | $T$(Jan '22) = 11.88 | $T$(Feb '22) = 10.25 | Feb '22 (-14%) | Nov '21 - Apr '22 |
| AIAN | Female | $T$(Jun '20) = 5.63 | $T$(Jul '20) = 6.96 | Jul '20 (+24%) | Apr-Sep '20 |
| AIAN | Female | $T$(Jan '22) = 7.06 | $T$(Feb '22) = 5.99 | Feb '22 (-15%) | Jan-Mar '22 |
| Black | Male | $T$(Feb '20) = 1.29 | $T$(Mar '20) = 1.46 | Mar '20 (+13%) | Dec '19 - May '20 |
| Black | Male | $T$(Jan '22) = 1.90 | $T$(Feb '22) = 1.56 | Feb '22 (-18%) | Jan-Mar '22 |
| Black | Female | $T$(Apr '20) = 0.50 | $T$(May '20) = 0.66 | May '20 (+33%) | Mar-Jun '20 |
| Black | Female | $T$(Jan '22) = 0.75 | $T$(Feb '22) = 0.57 | Feb '22 (-24%) | Jan-Mar '22 |
| Hispanic | Male | $T$(May '20) = 1.80 | $T$(Jun '20) = 2.11 | Jun '20 (+18%) | Mar-Jul '20 |
| Mixed race | Male | $T$(Apr '20) = 0.57 | $T$(May '20) = 0.75 | May '20 (+31%) | Nov '19 - Jul '20 |
| Mixed race | Male | $T$(Feb '22) = 0.74 | $T$(Mar '22) = 0.67 | Mar '22 (-10%) | Dec '21 - May '22 |
| White | Male | $T$(Mar '20) = 2.04 | $T$(Apr '20) = 2.48 | Apr '20 (+22%) | Mar-May '20 |
| White | Male | $T$(Jan '22) = 2.71 | $T$(Feb '22) = 2.48 | Feb '22 (-8%) | Jan-Apr '22 |
| White | Female | $T$(Mar '20) = 0.84 | $T$(Apr '20) = 0.94 | Apr '20 (+12%) | Mar-Jul '20 |

Values are plotted in Figure 3 of the main text. The percent change (pct chg) at month $t_i$ is defined as 100 $\mathrm{TCP}(t_i)\ /T(t_{i-1})$, where $\mathrm{TCP}(t_i) =\ T(t_i) - T(t_{i-1})$ is the TCP jump. Mortality is highest for the AIAN male and female populations. The largest relative increase is among AIAN males (TCP jump +42%, Jun '20) and among Black females (TCP jump +33%, May '20). Rbeast yields no TCP jumps for the Asian population.
